# A novel reporter system for temperature dependent analysis of nucleic acid amplification tests

**DOI:** 10.1101/2025.03.17.25324112

**Authors:** Dominik Nörz, Martin Aepfelbacher, Marc Lütgehetmann

## Abstract

**Background:** Real-time polymerase chain reaction (qPCR) is a key technology in molecular diagnostics, with multiplexing further improving efficiency for pathogen detection. However, multiplexing is limited by the number of optical channels available for probe differentiation. Temperature-dependent reporter systems like TOCE and TAGS address this by introducing temperature as a second analytical dimension, expanding multiplexing potential. Here, we introduce “Target-mediated branched overlap extension” (TBOE) as a new reporter technology that enables temperature-dependent signal generation without requiring probe digestion, making it compatible with both qPCR and isothermal amplification methods.

**Methods:** Prototype assays were designed using publicly available sequence data in conjunction with Geneious Prime and PrimerQuest software. The TBOE reporter consists of two oligonucleotide (Head and Tail oligo) that are positioned at two adjacent target sites within the amplified regions. The interaction between both oligos is facilitated by simultaneously binding to the target and a shared overlap sequence between the two oligos, thereby forming a triplex. The elongation of the head-oligo leads to duplex formation with the tag-sequence, thus generating detectable signals. Analytical experiments were carried out using clinical bacterial isolates containing carbapenemase genes or anonymized clinical remnant samples containing relevant bacterial target sequences.

**Results:** In contrast to TaqMan and TOCE, the TBOE reporter generated temperature dependent signals both in conjunction with 5’ exonuclease and without 5’ exonuclease activity in the mastermix used. A LAMP assay was created, demonstrating the ability of the TBOE reporter to generate temperature dependent signals in the context of an isothermal amplification assay. A set of carbapenemase assays were created to demonstrate the multiplexing potential of the TBOE reporter with both analysis by melt curve analysis and detection of different temperature levels during amplification. Finally, a SNP detection assay using TBOE reporters was created and tested in a clinical sample set.

**Discussion:** TaqMan-based multiplex qPCR has advanced pathogen detection but is limited in multiplexing capacity. We introduce Target-mediated branched overlap extension (TBOE), a novel temperature-dependent reporter system compatible with qPCR and isothermal amplification. TBOE enables high-order multiplexing without probe cleavage, expanding diagnostic applications, including SNP detection and LAMP-based assays.

## 1. Introduction

Real-time polymerase chain reaction (qPCR) allows for simultaneous amplification of a target nucleic acid through sequence-specific primers and quantification by detecting changes in fluorescence intensity during the reaction, generated by a reporter (1, 2). Early reporter systems like SYBR green and TaqMan probes remain widely used today and established qPCR as a cornerstone method in molecular biology (3, 4). Beyond the research lab, qPCR also revolutionized clinical microbiology by enabling highly sensitive and specific pathogen detection (5). In this context, qPCR multiplexing is particularly powerful, as it allows more efficient testing for a range of different pathogens while not requiring separate reactions for each individual one (6, 7). This approach has significantly improved diagnostic accuracy, turnaround time, cost efficiency, and patient management in infectious disease testing over the past decade (8).

Multiplexing potential for TaqMan based diagnostic qPCR tests is usually limited to 5-6 targets by the number of optical channels available on PCR instruments. To further expand the number of targets that can be covered in a single qPCR reaction, there has been an effort to develop reporter systems that are combinable with classic TaqMan probes, but also generate temperature dependent signals. One such example is the “tagged oligo cleavage and extension” (TOCE) method, which replaces the TaqMan probe with a 5’-tagged oligonucleotide, which is cleaved by a polymerase during the reaction and then serves as a primer to produce a counter-strand to a second reporter oligonucleotide (9). This method enables multiplexing well beyond the usual 5-6 targets (10). Another example of temperature dependent multiplexing is the “Temperature activated generation of signals” (TAGS) method, which uses a variation of a TaqMan probe, with an additional Taq- sequence, that can bind a second quencher oligonucleotide (patent: WO 2018/050828 A1). All of the mentioned methods are dependent on a polymerase featuring 5’ exonuclease activity in order to work, so they are not suitable for e.g. real-time loop mediated amplification (rt-LAMP), which is based on strand-displacement and cannot work in conjunction with 5’ exonuclease activity.

To overcome this limitation, we developed a new reporter technology: “Target-mediated branched overlap extension“ (TBOE). This new qPCR reporter system presented here generates temperature dependent signals and works in conjunction with classic TaqMan technology, but is not limited by the necessity to digest a probe or tagged oligonucleotide through 5’ exonuclease activity. The aim of the study was to and describe the new approach and prove the technology in different applications with potential for clinical diagnostics, including isothermal amplification.

## 2. Material and Methods

### 2.1 Assay design

Sequences for all respective targets were downloaded from Genbank and aligned using Geneious Prime software (Dotmatics, Boston, USA). Individual primer and probe sequences were generated either manually, or using PrimerQuest software (IDT, Coralville, USA). Primer sequences that are part of multiplex-tests were evaluated for interactions in-silico using Autodimerv1 software (11) and empirically by combining every primer with every other primer in a reaction containing intercalating dye (ResoLight, Roche, Switzerland) in either a matrix of water or random pooled clinical sample eluates.

All primer and reporter sequences are assembled in *supplementary material 1*.

Some primer and TBOE-head oligonucleotides include an internal 2’O-methly-RNA bases close to the 3’-end to delay the formation of primer dimers.

For TaqMan probes, oligonucleotides were labelled with a fluorophore (FAM, SUN, TexasRed, Atto647N or Cy5.5) at the 5’ end, and with either one dark-quencher at the 3’-end or at position 9 and a C3-Spacer at the 3’ end; or two dark quenchers, one at position 9 and one at the 3’-end (e.g. Zen/IBFQ).

TOCE (tagged oligo cleavage and extension) assays were designed according to published information (9). Briefly, a pitcher oligo was designed to bind to target between primer regions, and featuring a contrived tag sequence at its 5’ end. A further catcher oligo was designed, featuring the reverse complement of the pitcher-tag sequence at its 3’ end, and a contrived sequence with fluorophore and quencher pair at its 5’ end. The reporter was then operated with the same primer pair as the corresponding TaqMan and TBOE assays.

For TBOE reporter design, adjacent and conserved target regions were identified between the assays primer regions. For the first target region, a head-oligo was designed, featuring a target region at its 5’ end and a short overlap region on its 3’ end. For the second, downstream target region, a tail-oligo was designed, featuring a 5’ fluorescent tag sequence, including a fluorophore/quencher pair, approximately 18-22 bases apart; an overlap region, reverse complementary to the head-oligos overlap region, and a target region on its 3’ end and a C3-Spacer to avoid oligo elongation. In some cases, head and/or tail oligos may contain locked nucleic acids (LNA), to improve melting temperatures for respective sequence regions, or improve discrimination between sequence variations.

For TBOE capable LAMP assays, similar to regular LAMP design, three forward and three reverse target sequences were identified, to form a loop of 60-80 bases in length at one end of the dumbbell structure (12, 13). The TBOE reporter is fitted into the large dumbbell loop, whereas a loop primer in the small dumbbell loop.

### 2.2 NAAT reagents and run protocols

The following commercial PCR and LAMP mastermix formulations were used as part of this study: Quantabio (Beverly, USA) perfeCTa qPCR mastermix for TaqMan and general qPCR applications; Quantabio repliQa as an example of a mastermix featuring a polymerase without 5’ exonuclease activity and NEB Warmstart multi-purpose LAMP mastermix for LAMP applications (New England Biolabs, Ipswich, USA). To set up reactions, a 4x working stock of all required oligonucleotides was prepared and then combined with the mastermix reagents according to instructions by the manufacturer. Reactions volumes were 20µL total, 5µL nucleic acid eluate, unless indicated otherwise.

For each experiment, an identical copy of each PCR plate was created and experiments were then run on both a Bio-rad CFX96 and a Roche LightCycler-Pro. I.e. every experiment was repeated at least once.

Run protocols for individual experiments are available in *supplementary material 2*.

### 2.3 Dilution series and carbapenemase detection experiments

Carbapenemases are transferable, often plasmid borne beta-lactamases (“bla-“), which represent a growing problem for healthcare today (14). Detection of carbapenemase genes can be relevant in the context of screening within hospitals or for management of individual patients.

For bacterial isolate dilutions, a clinical isolate positive for the respective carbapenemase gene was inoculated in cobas PCR media (40% guanidine hydrochloride in Tris-HCl, Roche, Switzerland) and extracted using a MagNA-Pure 96 instrument (Roche). The nucleic acid eluate was then diluted in water to generate the required dilution series.

For multiplex carbapenemase detection experiments, eight clinical isolates positive for blaKPC, blaOXA-48, blaNDM, blaVIM, blaIMP, blaGES, blaOXA-23 and blaOXA-40/24 respectively, were retrieved from storage and processed as described above. The isolate positive for blaGES was also positive for blaVIM. Nucleic acid eluates were then diluted in water to a suitable concentration and used for experiments.

### 2.4 Clinical samples and gyrA Ser91Phe reference assay

The Ser91Phe substitution is a Quinolone-resistance marker in Neisseria gonorrhoeae (15). The detection of single-nucleotide polymorphisms is clinically relevant an many un-culturable or difficult to culture species, such as N. gonorrhoeae, Chlamydia sp. and Mycoplasma sp. (16).

Clinical samples were anonymized diagnostic remnant samples, either urethral, vaginal, rectal, throat swabs in amies medium (eSwab, copan, Italy). Samples were extracted using a MagNA-Pure 96 instrument. The Seegene Allplex NG + DR assay (Seegene, Seoul, South Korea) was run on these nucleic acid eluates to serve as a reference test. Results were then compared to the TBOE based Ser91Phe assay, which was performed twice, in a similar manner as described above.

The use of patient material was conducted in accordance with §12 of the Hamburg hospital law (§12 HmbKHG). The use of anonymized remnant diagnostic samples from patients was approved and informed consent was waived by the ethics committee of the Hamburg Medical Association (PV5626)

## 3. Results

### 3.1 Target-mediated branched overlap extension (TBOE)-assay concept

The novel “TBOE” reporter system consists of two independent oligonucleotides, one designated as “head-oligo” and the other as “tail-oligo”, see figure 1. Both have independent target regions to bind to a target nucleic acid, which are adjacent to each other (separated by 1-2 bases). Head- and tail-oligo carry target region sequences on their 5’- and 3’-ends respectively. The head-oligo is further able to bind to an overlap sequence on the tail-oligo, upstream of the target region, through a short overlap sequence on its 3’-end. By design, this low affinity interaction between head and tail oligo will only occur in the presence of target DNA. In addition to target- and overlap region, the tail-oligo has a fluorescent tag on its 5’ end, featuring a contrived sequence and a fluorophore-quencher pair, separated by 18-22 bases.

**Figure 1:**
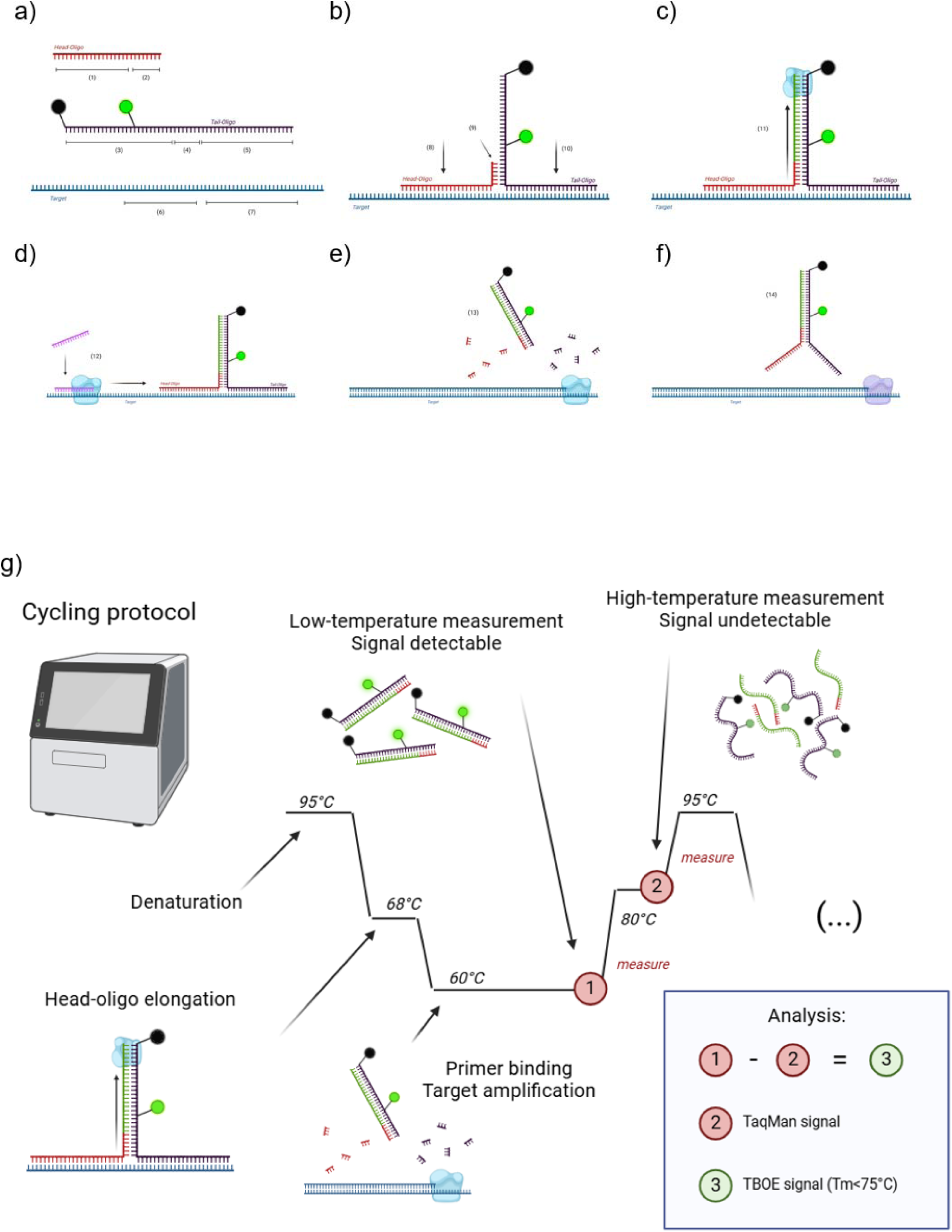
a-h: The TBOE reporter system consists of two independent oligonucleotides, one designated as “head-oligo” and the other as “tail-oligo”. The head oligo includes two different sequence regions, in 5’ to 3’ direction: The first sequence region of the head-oligo [1] binds to a target nucleic acid sequence. The second sequence [2] does not bind to target, but to a region on the tail-oligo [4]. The tail-oligo consists of a tag-sequence [3], containing fluorophore and quencher, an overlap region [4], which is reverse complementary to the corresponding overlap region of the head-oligo [2], and a target specific sequence region [5]. Head and tail-oligo bind to adjacent sequences on the target, [6] and [7]. Upon binding to a target nucleic acid, the head-oligo 3’ end hybridizes with the tail-oligo overlap region, [8] and [10], and enables generation of a counter-strand to the tag-sequence [9,11]. In a further step, temperature within the reaction is lowered to a point where regular PCR primers can bind and enable production of a counter-strand to the target [12]. In conjunction with polymerases featuring 5’ exonuclease activity, target specific regions within head-oligo [1] and tail-oligo [5] will be digested and the Tag-regions released [13]. In conjunction with polymerases not featuring 5’ exonuclease activity, the head/tail-complex will be displaced and released without cleavage [14]. g) Analysis through detection at different temperature levels: TBOE signals will become undetectable above the fluorescent tag sequence melting temperature. By detecting at two levels during amplification (1: below Tm and 2: well above Tm), the TBOE specific signals can be extracted by subtracting measurement 2 from measurement 1. (method similar to Lee et al.)

As mentioned, binding to a target sequence of both the head- and the tail-oligo allows interaction and enables synthesis of a new counter-strand to the fluorescent tag sequence by the polymerase. In consequence, the linearization of the tag sequence into double strand configuration will cause a rise in fluorescence due to the increased distance (ideally outside foster radius) between fluorophore and quencher, which results in a loss of the quenching effect of the dark-quencher.

Target sequences of head and tail oligo may be digested by a Taq-polymerase, or may be simply displaced by a polymerase without 5’ exonuclease activity as observed in isothermal amplification reactions. The functionality of the newly described reporter system is not impacted in either case. Moreover since the signal is dependent on the (reversible) duplex structure of the tail-oligo and the newly synthesised complementary stand, the reporter signal is temperature dependent (see Figure 1g and f). Furthermore, melt peaks of generated signals are determined by the contrived fluorescent tag sequence and thus not confounded by target sequence mutations.

### 3.2 TBOE signals are temperature dependent and do not require 5’ exonuclease activity

The beta-lactamase and carbapenemase gene “blaGES” is used as an exemplary target for this experiment.

In order to verify and compare the TBOE reporter concept with established qPCR reporter concepts, three assay variants for detection of blaGES featuring a TaqMan reporter, a TOCE reporter or a TBOE reporter, respectively, were combined with one mastermix containing a Taq-polymerase (Quanta perfecta), and a second mastermix containing a proofreading polymerase instead (Quantabio repliQa). Both versions of each assay were tested on a 10-fold dilution series of a blaGES positive clinical isolate. (Figure 2)

**Figure 2:**
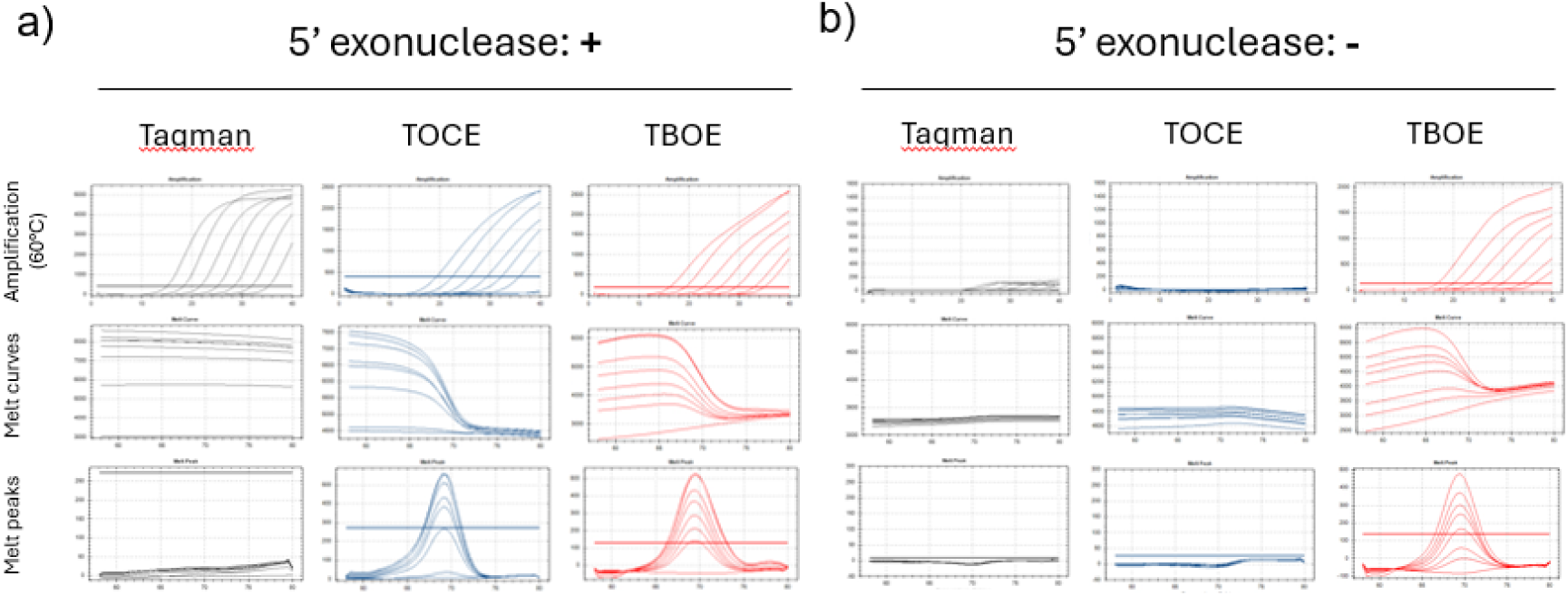
a) Comparing a TaqMan (black), TOCE (blue) and TBOE (red) based qPCR assay for detection of blaGES on a 10-fold dilution series. First row depicts amplification curves, y-axis: relative fluorescence units; x-axis: cycle. The second and third row depict melt curves and melt peaks respectively. y-axis: relative fluorescence units and negative delta of relative fluorescence units; x-axis: temperature. b) Same setup but using a polymerase without 5’ exonuclease detection (Quantabio repliQa HiFi mastermix).

Using the Taq-polymerase mastermix (figure 2a), all three assays yield detectable signals during amplification. For the TOCE (blue curves, middle) and TBOE assays (red curves, right), a melt-curve is detectable after amplification, whereas the TaqMan assay did not generate a melt-curves.

Using the HiFi-polymerase mastermix (figure 2b) without the 5′exonucleasae activity, the TaqMan reactions expectedly yield only very weak and abnormal amplification signals, whereas TOCE does not generate any signals at all. In contrast the TBOE assay generated strong signals and melt-curves, similar to what was observed in TaqMan mastermix.

### 3.3 TBOE works with rt-LAMP applications

To demonstrate the viability of a TBOE reporter in conjunction with isothermal amplification methods, a primer set for LAMP reactions, again targeting the blaGES gene, was created in such a way to incorporate the TBOE reporter system in one of the loop regions (similar to a loop primer). The assay was compared to the same primer set with high-resolution dye (ResoLight, Roche) using a 10-fold dilution series similar to 3.2 (Figure 3).

**Figure 3:**
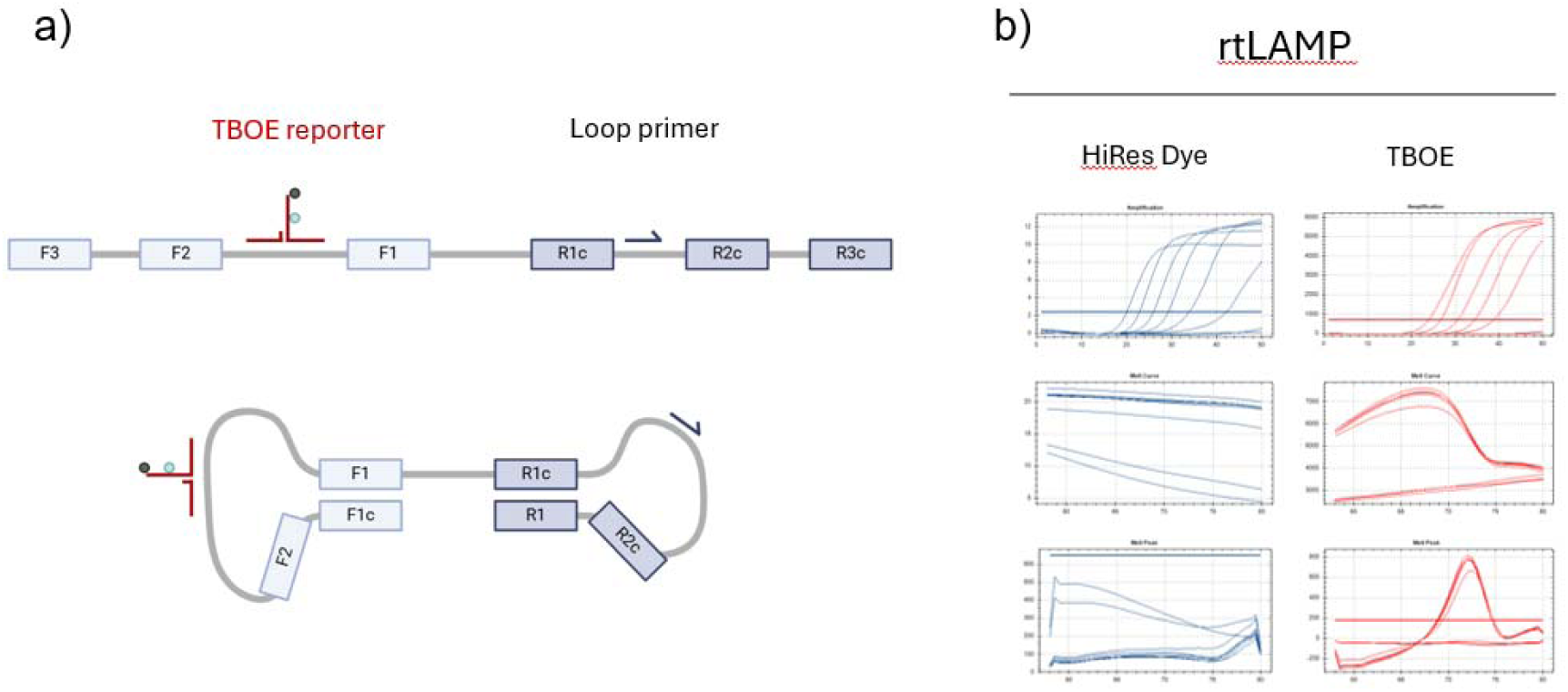
a) A classic LAMP design including a single loop primer was created to target blaGES, whereby one of the loop regions was made 60-80 bases long to incorporate the TBOE reporter. b) Amplification curves (first row), melt curves (second row) and melt peaks (third row) of a 10 fold dilution series of a blaGES positive Isolate. (graph setup similar to figure 2)

The HiRes dye version of the assay generated strong signals during amplification but no discernible melt-peaks. The TBOE-LAMP assay generated strong signals and melt-curves detectable after amplification (figure 3b).

### 3.4 Use of melt curve analysis to differentiate different assays within a multiplex test

The carbapenemase genes blaKPC, blaOXA-48, blaNDM, blaVIM, blaIMP, blaGES, blaOXA-23 and bla- OXA40/24 are among the most important genes for carbapenem resistance in gram-negative bacteria in human disease (17, 18).

To demonstrate the practical viability of TBOE based multiplexing, a multiplex test incorporating 8 different TBOE-assays was assembled to allow differentiation via melt curve analysis and target specific Tm on four different optical channels. As proof-of-concept, predetermined samples with all 8 targets were run in triplicates to verify assay functionality (Figure 4). (The isolate positive for blaGES was also positive for blaVIM)

**Figure 4:**
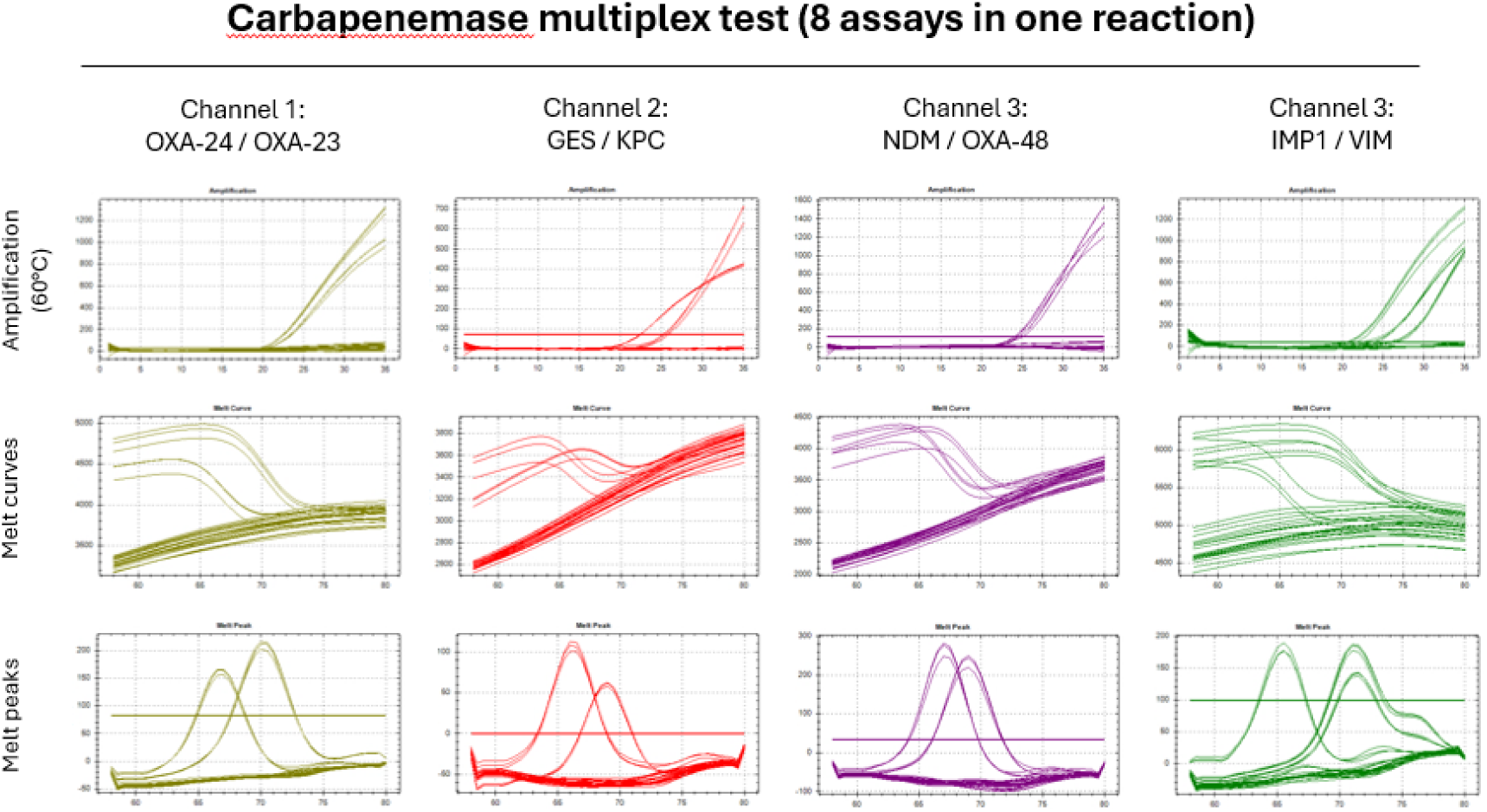
Amplification curves, melt curves and melt peaks of each individual channel for the carbapenemase multiplex test (graph setup similar to figure 2). Each target is allocated to one detection channel and one individual Tm. Melt peak analysis allows differentiation of individual targets.

Results were called through melt-peak analysis, whereby each peak above threshold constitutes a positive result for the allocated target. All eight samples and negative control were correctly detected by the multiplex test with amplification curves and melt curves in the correct optical channels (Figure 4, table 1) .

**Table 1:** Melt peak analysis of all eight samples and negative control. Samples were run in triplicates; indicated melting temperatures are averages. Melt peak analysis was carried out on a LC-Pro instrument.

| Sample | contains | Ch2: JOE | Ch3: TEX | Ch4: 647 | Ch5: Cy5.5 |
| --- | --- | --- | --- | --- | --- |
| 1 | blaOXA-23 | 67.3 |  |  |  |
| 2 | blaOXA-40/24 | 70.8 |  |  |  |
| 3 | blaKPC |  | 66.7 |  |  |
| 4 | blaGES, blaVIM |  | 69.6 |  | 71.8 |
| 5 | blaOXA-48 |  |  | 69.5 |  |
| 6 | blaNDM |  |  | 67.7 |  |
| 7 | blaVIM |  |  |  | 72 |
| 8 | blaIMP |  |  |  | 66 |
| neg | - |  |  |  |  |

### 3.5 qPCR amplification curve reconstruction from fluorescence detection at multiple temperature levels in a multiplex of both TBOE and TaqMan assays

A separate set of assays for blaVIM, blaNDM and blaOXA-48-like was used to demonstrate assay differentiation through different temperature levels during qPCR amplification. blaOXA-48 and blaNDM assays were designed to be undetectable at 72°C and 90°C respectively. For blaVIM, a TaqMan assay was used (not temperature dependent). The assays were run on every possible combination of targets in triplicates to verify functionality (Figure 4).

In a first step, amplification signals are detected during amplification at 60°C (step 4), 72°C (step 5) and 90°C (step 6) (figure 5a). Target specific signals were extracted from the raw data by compensating between the different steps as shown in figure 5b. Compensated signals are unambiguous for the respective targets (figure 5c) and a deconflicted table of end-point fluorescence showed a perfect match to the experiment setup (figure 5d).

**Figure 5:**
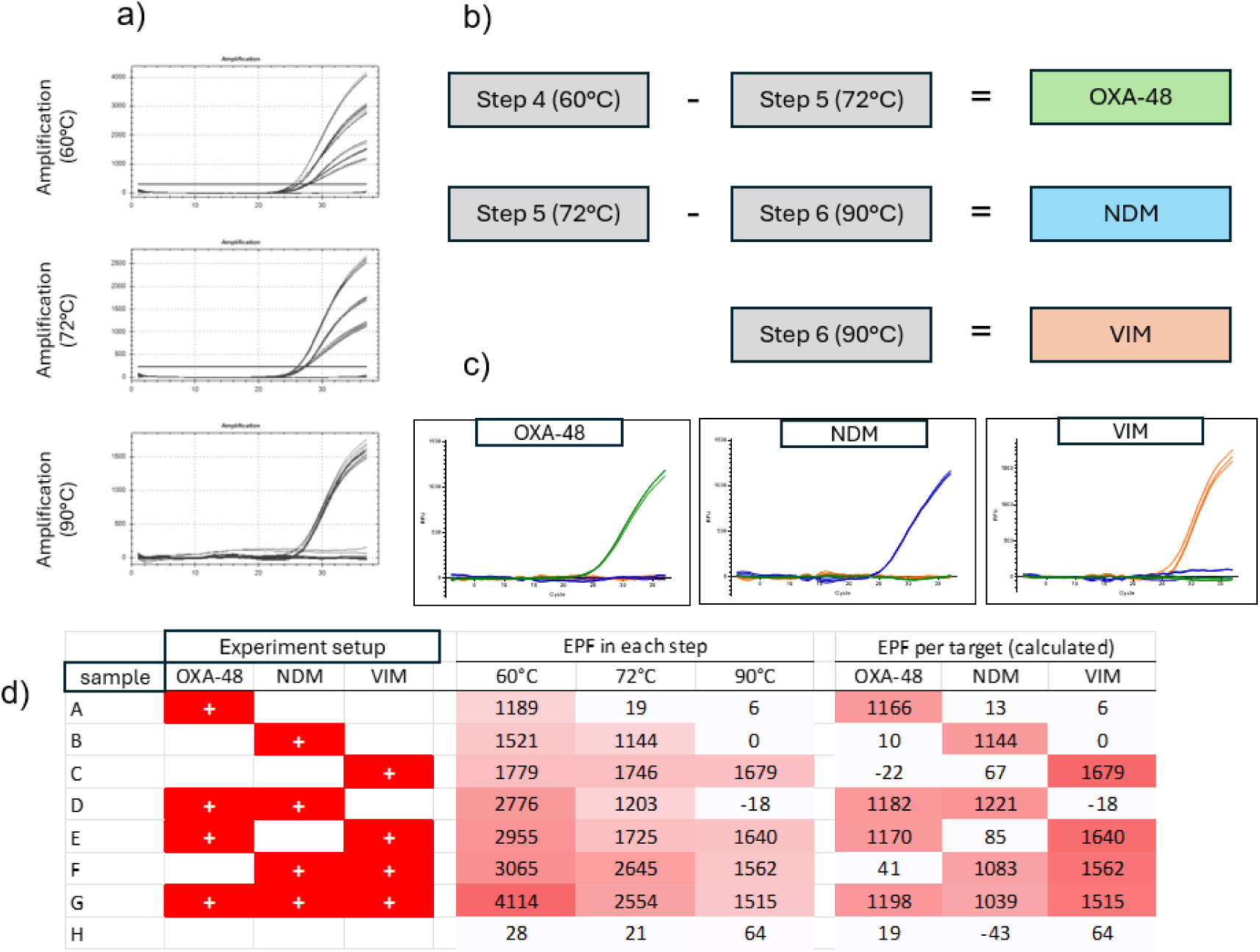
Signal deconfliction through detection at different temperatures during amplification. a) shows raw curves, detected at 60°C, 72°C and 90°C. b) Individual target signals are extracted through subtraction (including correction factor) of raw RFU readings of individual temperature steps. c) Individual amplification curves can be reconstructed by inputting calculated RFUs into a new graph (Graphpad prism). d) Experiment setup compared to end-point-fluorescence (EPF) of each raw temperature step and calculated EPF of signals after deconfliction. The calculated EPF values match the experimental setup of each sample.

### 3.6 Detection of a single nucleotide polymorphism using competitive TBOE tail oligonucleotides

In order to demonstrate the possibility to discriminate single-nucleotide polymorphisms (SNP) using a TBOE assay, a Neisseria gonorrhoeae gyrA Ser91Phe (Quinolone resistance marker) assay was created, using a set of two competitive TBOE tails, one for each sequence variation. LNA was used to increase sequence specificity for the tail-oligos target regions. A total of 37 clinical swab samples were included in the experiment (eSwab, copan, urethral, anal, throat, vaginal). Clinical samples were predetermined positive or negative using a cobas5800 based qPCR screening assay (Roche, Rotkreutz, Switzerland). Of these samples, 16 were negative and 21 positive for N. gonorrhoeae DNA. All samples were then run on the TBOE gyrA Ser91Phe and the commercial Seegene Allplex NG + DR assay.

To distinguish the two relevant sequence variations using the TBOE assay, melt curve analysis was carried out, whereby a maximum peak at approximately 66.5°C indicated the wild type sequence, and a maximum peak at approximately 72°C indicated the mutated sequence for Ser91Phe (Quinolone resistant) (see figure 6).

**Figure 6:**
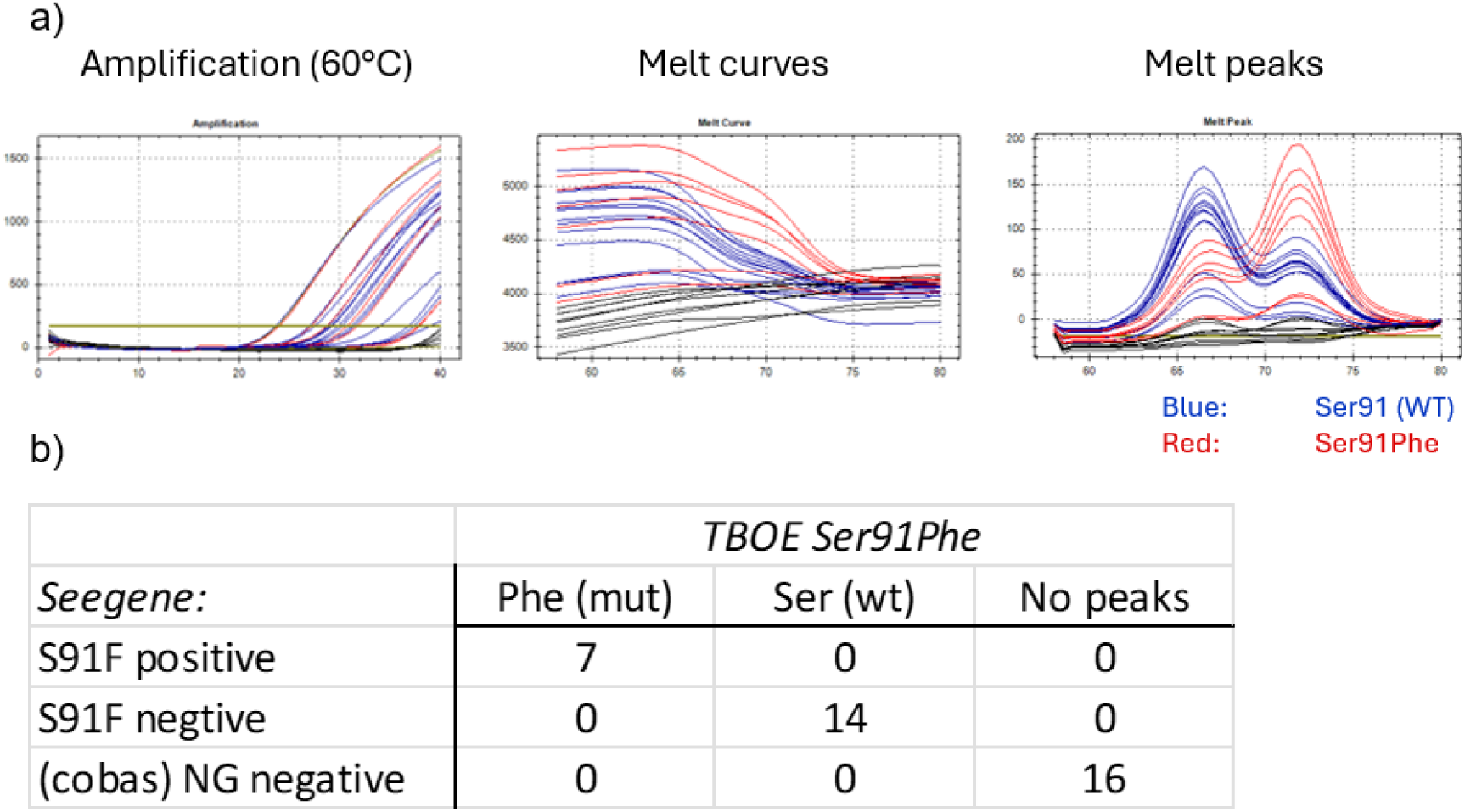
a) Amplification curves of clinical samples run on the TBOE Ser91Phe assay. Top: amplification curves, middle: melt curves, bottom: melt peaks. Blue curves indicate wild type sequence (melt peak at 66.5°C) and red curves indicate Ser91Phe substitution (melt peak at 72°C). b) Results of clinical samples compared for this experiment. 16 samples were pre-determined negative for N. gonorrhoeae and did not yield melt peaks. 14 samples were positive for N. gonorrhoeae and negative for Ser91Phe in the reference assay (Seegene Allplex NG + DR). 7 Samples were positive for Ser91Phe in the reference assay. The TBOE based assay identified all sequence variations correctly.

All samples featuring a peak at 72°C returned a positive result for Ser91Phe on the Seegene reference assay and all samples featuring a peak at 66.5°C returned a negative result on the Seegene reference assay. I.e. positive and negative agreement were 100% for the detection of the Ser91Phe SNP (figure 6, supplementary material 3).

## 4. Discussion and Conclusion

TaqMan-based multiplex-PCR has had a significant impact on sensitivity, specificity and efficiency of pathogen detection in clinical microbiology over the past two decades (6–8). More recently, syndromic panels have further broadened diagnostic options and there is increasing evidence for tangible clinical benefits such as reduced length of stay, reduced antibiotics use and better allocation of isolation capacity, e.g. in the context of respiratory virus panels or gastroenteritis panels (19–21). Furthermore, PCR panels are increasingly used for positive blood culture identification and have been shown to enhance antibiotic stewardship programs by enabling rapid pathogen identification and detection of resistance genes (22). However, detecting the large number of targets required (typically more than 20) renders conventional low-order multiplexing impractical for these applications, thus requiring novel methods for higher-order multiplexing.

In this study, we demonstrated and compared the novel “target-mediated branched overlap extension” (TBOE) reporter system with qPCR reporters already established in the field of molecular diagnostics. We showed that TBOE amplification signals are temperature dependent and can generate melt-curves, similar to the “tagged oligo cleavage and extension” (TOCE) method (9). In contrast to TOCE and the “temperature activated generation of signals” (TAGS) method however, TBOE does not require cleavage of any of its components in order to generate signals. Therefore it was possible to generate strong signals with polymerases that do not have 5’ exonuclease activity, such as in the repliQa mastermix (Quantabio), or the Bst 2.0 polymerase for LAMP applications, which does not work with any of the afore mentioned methods. The potential value for diagnostic applications was further demonstrated by a proof-of-concept TBOE based 8-plex assay for detection of 8 different carbapenemase genes in a single reaction and melt curve analysis in four optical channels. Furthermore, we showed that individual qPCR amplification curves can be extracted from raw data by measuring fluorescence at multiple temperature levels during amplification, similar to the “Multiple Detection Temperatures” (MuDT) or the TAGS concept by Roche (9). Finally, we demonstrated the viability of the TBOE method for detection of single-nucleotide polymorphisms in a target commonly used in clinical diagnostics.

Besides TOCE and TAGS, other temperature dependent qPCR reporter systems have been published over the years, such as molecular beacons (23), scorpion primers (24), FRET hybridization probes (25) and others (26). In contrast to TOCE, TAGS and TBOE, these systems are often incompatible with 5’ exonuclease polymerases, as important functional parts of the reporter would be digested. This precludes applications where such reporters may be combined with TaqMan assays in order to expand multiplexing potential. Their melting-temperature is also in many cases dependent on the target-sequence, thus making them vulnerable to Tm differences due to mutations.

On the other hand, methods like TOCE and TAGS, in turn, are not suitable for applications like LAMP, where 5’ exonuclease activity can not be part of the reaction due to the nature of the amplification process.

In conclusion, we have demonstrated the functionality of a novel, temperature dependent qPCR reporter system for high-multiplexing applications. TBOE can be combined with TaqMan assays, similar to TOCE and TAGS, but can also be used for temperature dependent detection of LAMP reactions, as it does not requiring cleavage in order to work. When using a thermocycler with five optical channels, up to 15 different targets is feasible through either melt-curve based analysis or detection of amplification at different temperature levels, as demonstrated in this study.

## Data Availability

All data produced in the present study are available upon reasonable request to the authors

## Acknowledgments

None

## 6. Abbreviations

Dcp, digital copies; LoD, Limit of Detection; IC, internal control; IVD, in-vitro diagnostic; RFI, relative fluorescence increase; CI, confidence interval;

## 7. Author contribution

DN, MA and ML conceptualized and supervised the study. DN performed the experiments. DN, MA and ML wrote and edited the manuscript. All authors agreed to the publication of the final manuscript.

## References

1. Mullis KB, Faloona FA. [21] Specific synthesis of DNA in vitro via a polymerase-catalyzed chain reaction. Methods in Enzymology. 155: Academic Press; 1987. p. 335-50.

2. 2. Higuchi R, Fockler C, Dollinger G, Watson R. Kinetic PCR analysis: real-time monitoring of DNA amplification reactions. Bio/technology (Nature Publishing Company). 1993;11(9):1026-30.

3. Heid CA, Stevens J, Livak KJ, Williams PM. Real time quantitative PCR. Genome research. 1996;6(10):986–94.

4. Zipper H, Brunner H, Bernhagen J, Vitzthum F. Investigations on DNA intercalation and surface binding by SYBR Green I, its structure determination and methodological implications. Nucleic acids research. 2004;32(12):e103.

5. Espy MJ, Uhl JR, Sloan LM, Buckwalter SP, Jones MF, Vetter EA, et al. Real-time PCR in clinical microbiology: applications for routine laboratory testing. Clinical microbiology reviews. 2006;19(1):165–256.

6. Gunson RN, Collins TC, Carman WF. Real-time RT-PCR detection of 12 respiratory viral infections in four triplex reactions. Journal of clinical virology : the official publication of the Pan American Society for Clinical Virology. 2005;33(4):341–4.

7. Kuypers J, Wright N, Ferrenberg J, Huang M-L, Cent A, Corey L, et al. Comparison of Real-Time PCR Assays with Fluorescent-Antibody Assays for Diagnosis of Respiratory Virus Infections in Children. Journal of Clinical Microbiology. 2006;44(7):2382–8.

8. Hanson KE, Couturier MR. Multiplexed Molecular Diagnostics for Respiratory, Gastrointestinal, and Central Nervous System Infections. Clinical infectious diseases : an official publication of the Infectious Diseases Society of America. 2016;63(10):1361–7.

9. Lee Y-J, Kim D, Lee K, Chun J-Y. Single-channel multiplexing without melting curve analysis in real-time PCR. Scientific Reports. 2014;4(1):7439.

10. Cho CH, Chulten B, Lee CK, Nam MH, Yoon SY, Lim CS, et al. Evaluation of a novel real-time RT-PCR using TOCE technology compared with culture and Seeplex RV15 for simultaneous detection of respiratory viruses. Journal of clinical virology : the official publication of the Pan American Society for Clinical Virology. 2013;57(4):338–42.

11. Vallone PM, Butler JM. AutoDimer: a screening tool for primer-dimer and hairpin structures. BioTechniques. 2004;37(2):226–31.

12. Notomi T, Okayama H, Masubuchi H, Yonekawa T, Watanabe K, Amino N, et al. Loop-mediated isothermal amplification of DNA. Nucleic acids research. 2000;28(12):E63.

13. Nagamine K, Hase T, Notomi T. Accelerated reaction by loop-mediated isothermal amplification using loop primers. Molecular and Cellular Probes. 2002;16(3):223–9.

14. Naghavi M, Vollset SE, Ikuta KS, Swetschinski LR, Gray AP, Wool EE, et al. Global burden of bacterial antimicrobial resistance 1990&#x2013;2021: a systematic analysis with forecasts to 2050. The Lancet. 2024;404(10459):1199–226.

15. Tanaka M, Takahashi K, Saika T, Kobayashi I, Ueno T, Kumazawa J. Development of fluoroquinolone resistance and mutations involving GyrA and ParC proteins among Neisseria gonorrhoeae isolates in Japan. J Urol. 1998;159(6):2215–9.

16. Twin J, Jensen JS, Bradshaw CS, Garland SM, Fairley CK, Min LY, et al. Transmission and selection of macrolide resistant Mycoplasma genitalium infections detected by rapid high resolution melt analysis. PLoS One. 2012;7(4):e35593.

17. Bonnin RA, Jousset AB, Emeraud C, Oueslati S, Dortet L, Naas T. Genetic Diversity, Biochemical Properties, and Detection Methods of Minor Carbapenemases in Enterobacterales. 2021;7.

18. Perez F, Hujer Andrea M, Hujer Kristine M, Decker Brooke K, Rather Philip N, Bonomo Robert A. Global Challenge of Multidrug-Resistant Acinetobacter baumannii. Antimicrobial Agents and Chemotherapy. 2007;51(10):3471–84.

19. Clark TW, Lindsley K, Wigmosta TB, Bhagat A, Hemmert RB, Uyei J, et al. Rapid multiplex PCR for respiratory viruses reduces time to result and improves clinical care: Results of a systematic review and meta-analysis. Journal of Infection. 2023;86(5):462–75.

20. Brendish NJ, Beard KR, Malachira AK, Tanner AR, Sanga-Nyirongo L, Gwiggner M, et al. Clinical impact of syndromic molecular point-of-care testing for gastrointestinal pathogens in adults hospitalised with suspected gastroenteritis (GastroPOC): a pragmatic, open-label, randomised controlled trial. The Lancet Infectious Diseases. 2023;23(8):945–55.

21. Lewinski MA, Alby K, Babady NE, Butler-Wu SM, Bard JD, Greninger AL, et al. Exploring the Utility of Multiplex Infectious Disease Panel Testing for Diagnosis of Infection in Different Body Sites: A Joint Report of the Association for Molecular Pathology, American Society for Microbiology, Infectious Diseases Society of America, and Pan American Society for Clinical Virology. The Journal of Molecular Diagnostics. 2023;25(12):857–75.

22. MacVane Shawn H, Nolte Frederick S. Benefits of Adding a Rapid PCR-Based Blood Culture Identification Panel to an Established Antimicrobial Stewardship Program. Journal of Clinical Microbiology. 2016;54(10):2455–63.

23. Tyagi S, Kramer FR. Molecular beacons: probes that fluoresce upon hybridization. Nature biotechnology. 1996;14(3):303–8.

24. Thelwell N, Millington S, Solinas A, Booth J, Brown T. Mode of action and application of Scorpion primers to mutation detection. Nucleic acids research. 2000;28(19):3752–61.

25. Didenko VV. DNA probes using fluorescence resonance energy transfer (FRET): designs and applications. BioTechniques. 2001;31(5):1106–16, 18, 20-1.

26. Marras SAE, Tyagi S, Kramer FR. Real-time assays with molecular beacons and other fluorescent nucleic acid hybridization probes. Clinica Chimica Acta. 2006;363(1):48–60.

