## Supplementary material for "A novel reporter system for temperature dependent analysis of nucleic acid amplification tests"

### Supplemental material 1: Oligo sequences

Part 1

| **blaGES (Taqman)** | Oligo sequence (5' - 3') |
|---|---|
| GES-fwd | CTGTGGCTAAAGTCCTCTATG |
| GES-rev | CAACAACCCAATCTTTAGGAAA |
| GES-Probe | FAM- CGT CTC CCG (ZEN)TTT GGT TTC CG - IBFQ |
| **blaGES (TOCE)** | Oligo sequence (5' - 3') |
| GES-fwd | CTGTGGCTAAAGTCCTCTATG |
| GES-rev | CAACAACCCAATCTTTAGGAAA |
| AR1-GES-tc1-P1 | GGT CCT TCA TCG CTC CGT CTC CCG TTT GGT TTC CG -C3-Spacer |
| AR1-GES-tc1-C1 | 5'BHQ2- TTT TTT TTT TTT TTT TC(FAM-dT) CTC TTG ACG GAG CGA TGA AGG ACC -C3-Spacer |
| **blaGES (TBOE)** | Oligo sequence (5' - 3') |
| v3GES-Fv2 | AACACACCTGGCGACCT |
| v3GES-Rv4 | CAATCTTTAGGAAAACCCGC |
| v3GES-H1-a | GTCGCGTCTCCCGTTTGGTTT T AT GAC TCT |
| v3GES-T1-a | 5'BHQ2- AAA CAG CCC GCG GTC GTT T(TexasRed-dT)C CTT TT AGA GTC AT T CGATCAGCCACCTCTCAATGGTGTGGGT -C3-Spacer |

| **blaGES (TBOE-LAMP)** | Oligo sequence (5' - 3') |
|---|---|
| v3GES-L-F2L | TTAGGAAAACCCGCTCGTA TTT GCGCACTGACGTCCAC |
| v3GES-L-F3 | CGTACTGTGGCTAAAGTCCT |
| v3GES-L-R2L | CCTGCGCCAACGGG TTT GTAATCTCTCTCCTGGGCTT |
| v3GES-L-R3 | GCCGTTGTATACACCGCT |
| v3GES-L-RLP | CCGGAACGACATTGGTTT |
| v3GES-H1-a | GTCGCGTCTCCCGTTTGGTTT T AT GAC TCT |
| v3GES-T1-a | 5'BHQ2- AAA CAG CCC GCG GTC GTT T(TexasRed-dT)C CTT TT AGA GTC AT T CGATCAGCCACCTCTCAATGGTGTGGGT -C3-Spacer |

### Supplemental material 1: Oligo sequences

Part 2

| **AR1C-X1** | Oligo sequence (5' - 3') |
|---|---|
| v3IMP-br1-F1 | AGA GBC TTT GCC AGA TTT AAA AAT TG |
| v3IMP-br1-R1 | TAC AAG AAC MAC CAA ACC ATG TTT |
| v3IMP-br1-R2 | AGA ACM ACC AAG CCG TGT TT |
| v3VIM-F1 | CCR TCC AAT GGT CTC ATT GTC C |
| v3VIM-F2 | TCG TCT AAT GGR CTT ATC GTC C |
| v3VIM-R1 | GCC GAC GCG GTC GTC AT |
| v3VIM-R2 | CCA CCG ACT CGA TCG TCA T |
| v3NDM-F1 | GCT GGC GGT GGT GAC TC |
| v3NDM-Ralt1 | GGT TGC TGG TTC GAC CC |
| v3_2_OX48-F1 | AGCCTTATCGGCTGTGTTT |
| v3_2_OX48-R1 | TTTATGTTCAGTAAAGTGAGCATTCC |
| v3KPC-F1 | CGC TAA ACT CGA ACA GGA CTT T |
| v3KPC-R1 | AGT GGG AAG CGC TCC TC |
| v3GES-Fv2 | AACACACCTGGCGACCT |
| v3GES-Rv4 | CAATCTTTAGGAAAACCCGC |
| v3OXA23-F1 | CTACATTTAAAATGTTGAATGCCCT |
| v3OXA23-R1 | ATTACCGAAAYCAATACGTTYTACTTCTT |
| v3OXA24-F1 | TGGCATTGTCAGCAGTTCC |
| v3OXA24-R1 | CCAACTAACCAAAAATTATCGACCT |
| v3IMP-br1-H1-a | ACR CCC CAC CCG TTA ACT TCY T A AT TGC TCT |
| v3IMP-br1-T1-a | 5'BHQ2- AAA GTG CCG GTC TTT TTT CT(Cy5.5-dT) TTC TT AGA GCA AT A AAA CGA AGT ATG AAC ATA AAC GCC TTC RTC AAG -C3-Spacer |
| v3VIM-H1-b | G+CM C+TT CTC GCG GAG ATT GA T TA TGG TCT |
| v3VIM-T1-a | 5'BHQ2- AAA CTC GCC GCG GTC CTC CT(Cy5.5-dT) TTT CT AGA CCA TA T AAG CAA ATT GGA CTT CCY GTA ACG CG -C3-Spacer |
| v3NDM-H1-a | GCC GCA TGC AGC GCG T A TA CTG TCT |
| v3NDM-T1-a | 5'BHQ2- AAA GTC GCG GTG CTT TTT TC(Atto647N-dT) TTC TT AGA CAG TA T CAT ACC GCC CAT CTT GTC CTG ATG CG -C3-Spacer |
| v3_2_OX48-H1-a | TTCTTGCCATTCCTTTGCTACCGC C AT GGT TCT |
| v3_2_OX48-T1-a | 5'BHQ2- AAA CAC GCC CCG GTG TTC CT(Atto647N-dT) CTT TT AGA ACC AT C GGCATTCCGATAATCGATGCCACC -C3-Spacer |
| v3KPC-H1-a | CCA TCG GTG TGT ACG CGA TGG AT T TA GTC TCT |
| v3KPC-T1-a | 5'BHQ2- AAA GAC GCC GTG CTT TTT C(TexasRed-dT)T TCT TT AGA GAC TA T CCG GCT CAG GCG CAA CTG TAA GT -C3-Spacer |
| v3GES-H1-a | GTCGCGTCTCCCGTTTGGTTT T AT GAC TCT |
| v3GES-T1-a | 5'BHQ2- AAA CAG CCC GCG GTC GTT T(TexasRed-dT)C CTT TT AGA GTC AT T CGATCAGCCACCTCTCAATGGTGTGGGT -C3-Spacer |
| v3OXA23-H1-a | GAAAAAGACATGACACTAGGAGAAGCCATGA T TA GCT CTT |
| v3OXA23-T1-a | 5'BHQ2- AAA GTG CCC GTC CTT TTT (JOE-dT)TC CTT TT AAG AGC TA T GCTTTCYGCAGTCCCAGTCTATCAGGAAC -C3-Spacer |
| v3OXA24-H1-a | GTATTTCCAAAATTAACCCGCTTTACTTCTTTC A TA CAG TCT |
| v3OXA24-T1-a | 5'BHQ2- AAA CTG CCG CCC GTG CTT (JOE-dT)CT TCT TT AGA CTG TA A GCATTAGCTCTAGGCCAGTCCGTCTTGC -C3-Spacer |

# Supplemental material 1: Oligo sequences                Part 3

| **BEX-Joe** | Oligo sequence (5' - 3') |
|---|---|
| OXA48-Fwd | AGGGCGTAGTTGTGC(O-meth-U)C |
| OXA48-Rev | GTGTTCATCCTTAACCAC(O-meth-G)C |
| NDM-Fwd | GCCACACCAGTGACAATA(O-meth-U)C |
| NDM-Rev | GTGCTCAGTGTCGG(O-meth-C)AT |
| v3VIM-F1 | CCR TCC AAT GGT CTC ATT GTC C |
| v3VIM-F2 | TCG TCT AAT GGR CTT ATC GTC C |
| v3VIM-R1 | GCC GAC GCG GTC GTC AT |
| v3VIM-R2 | CCA CCG ACT CGA TCG TCA T |
| Oxa48-be1-head1 | GAA TAA GCA GCA AGG ATT TAC CAA TAA TCT T TTT CCT ACG |
| Oxa48-be1-tail1 | 5'BHQ2- CGT TCC GTC TTT TTT TTT TC(JOE-dT) TCT T CGT AGG AAT AAC GGG CGA ACC AAG CAT TTT TAC C -C3-Spacer |
| NDM-be2-head1 | GCG ACT TGG CCT TGC TGT AAA CCT GAG |
| NDM-be2-tail1 | 5'BHQ2- GCC GTG CTC GTC GTC CTC CT(JOE-dT) CTC T CTC AGG TTA CTT GAT CAG GCA GCC ACC AAA AGC G -C3-Spacer |
| VIM-P | SUN- ATGAGTTGC(ZEN)TTTTGATTGATACAGCKTGG -IBFQ |
| **SC2-mc** | Oligo sequence (5' - 3') |
| RpT-SC2-mc-F1 | TGGTTGTTAATGCAGCCAATGTTT |
| RpT-SC2-mc-R1 | GGTCCATTAGTAGCTATGTAATCATC |
| RpT-SC2-mc-H1-a | AACTTGCATGGCATTGTTAGTAGCCTTATTTA T AT GAC TCT |
| RpT-SC2-mc-T1-a | BHQ2- TAA CAG CCC GCG GTC GTG TC(Cy5.5-dT) TCT TTC TT AGA GTC AT T GGCTCCTGCAACACCTCCTCCATG -C3-Spacer |
| **NG Ser91Phe** | Oligo sequence (5' - 3') |
| NG-gyra-91-F | CTGGAATGCCGCCTACAA |
| NG-gyra-91-R | GTTGCCCTGTCCGTCTATC |
| NG-gyra-91-H1-b | CGGTAAATACCACCCCCACGGC T TA TGC TCT A |
| NG-gyra-91ser-T1-a | BHQ2- AAA GTG CCG GTC TTT TTT C(JOE-dT)T TTC TT AGA GCA TA T ATT +CCG CAG TTT ACG VCA CCA T -C3-Spacer |
| NG-gyra-91phe-T1-a | BHQ2- AAA CTC GCC GCG GTC CTC C(JOE-dT)T TTT CT AGA GCA TA T ATT +TCG CAG TTT ACG VCA CCA TC -C3-Spacer |

### Supplemental material 2: Run protocols

| Quanta_perfeCTa_TBOE_generic | | | |
|---|---|---|---|
| Step | Temp. | Duration | Operation |
| 1 | 95°C | 5:00 | Hold |
| 2 | 95°C | 0:10 | |
| 3 | 68°C | 0:05 | |
| 4 | 60°C | 0:25 | |
| | +Detect | | Cycle |
| 5 | 80°C | 0:05 | |
| | +Detect | | |
| | go to Step 2: 39x | | |
| 6 | 58°C | 0:30 | Hold |
| 7 | 85°C | continuous | Melt |

| Quanta_repliQa_TBOE | | | |
|---|---|---|---|
| Step | Temp. | Duration | Operation |
| 1 | 98°C | 0:30 | Hold |
| 2 | 98°C | 0:10 | |
| 3 | 68°C | 0:05 | |
| 4 | 60°C | 0:20 | |
| | +Detect | | Cycle |
| 5 | 80°C | 0:05 | |
| | +Detect | | |
| | go to Step 2: 39x | | |
| 6 | 58°C | 0:30 | Hold |
| 7 | 85°C | continuous | Melt |

| NEB WarmStart TBOE | | | |
|---|---|---|---|
| Step | Temp. | Duration | Operation |
| 1 | 65°C | 0:30 | Hold |
| 2 | 60°C | 0:30 | |
| | +Detect | | Cycle |
| | go to Step 2: 49x | | |
| 3 | 95°C | 0:30 | |
| 4 | 55°C | 0:30 | Hold |
| 5 | 85°C | continuous | Melt |

| Quanta_perfeCTa_BEX | | | |
|---|---|---|---|
| Step | Temp. | Duration | Operation |
| 1 | 95°C | 5:00 | Hold |
| 2 | 95°C | 0:10 | |
| 3 | 68°C | 0:05 | |
| 4 | 60°C | 0:25 | |
| | +Detect | | |
| 5 | 72°C | 0:05 | Cycle |
| | +Detect | | |
| 6 | 90°C | 0:05 | |
| | +Detect | | |
| | go to Step 2: 39x | | |
| 7 | 58°C | 0:30 | Hold |
| 8 | 85°C | continuous | Melt |

| Quanta_ULTRAplex_TBOE_generic | | | |
|---|---|---|---|
| Step | Temp. | Duration | Operation |
| 1 | 54°C | 10:00 | |
| 2 | 95°C | 3:00 | Hold |
| 3 | 95°C | 0:05 | |
| 4 | 68°C | 0:05 | |
| 5 | 60°C | 0:25 | |
| | +Detect | | Cycle |
| 6 | 80°C | 0:05 | |
| | +Detect | | |
| | go to Step 2: 39x | | |
| 7 | 58°C | 0:30 | Hold |
| 8 | 85°C | continuous | Melt |

### Supplemental material 3: Clinical set, N. gonorrhoeae Ser91Phe

| Material (swab) | cobas NG Tgt1 | TBOE ct | Melt peak | Interpretation | SG: NG1 | SG: Ser91Phe |
|---|---|---|---|---|---|---|
| Urin | 18.9 | 26.6 | 72 | ***Phe*** | 23.4 | ***26.3*** |
| Vaginal | 19.3 | 23.4 | 72 | ***Phe*** | 20.3 | ***23.1*** |
| Urethra | 20.8 | 28.1 | 72 | ***Phe*** | 25.6 | ***28.7*** |
| Anal | 23.3 | 30.2 | 72 | ***Phe*** | 27 | ***29.9*** |
| Anal | 24.5 | 31.2 | 72 | ***Phe*** | 28.4 | ***31.5*** |
| Anal | 30.2 | 37.3 | 72 | ***Phe*** | 35.3 | ***38.5*** |
| Throat | 31 | 37.2 | 72 | ***Phe*** | 35.6 | ***40.1*** |
| Urethra | 20 | 23.1 | 66.5 | Ser | 19.8 | |
| Urethra | 20.6 | 28.6 | 66.5 | Ser | 25.9 | |
| Urethra | 20.6 | 28.5 | 66.5 | Ser | 25.9 | |
| Urethra | 21 | 26.5 | 66.5 | Ser | 23.1 | |
| Anal | 21.6 | 29.3 | 66.5 | Ser | 26.2 | |
| Anal | 21.6 | 26.9 | 66.5 | Ser | 24.1 | |
| Anal | 21.6 | 28.3 | 66.5 | Ser | 25.1 | |
| Anal | 22.9 | 30.6 | 66.5 | Ser | 27.4 | |
| Throat | 23.5 | 31 | 66.5 | Ser | 27.9 | |
| Anal | 24.2 | 29.7 | 66.5 | Ser | 26.6 | |
| Throat | 25.7 | 33.3 | 66.5 | Ser | 30.5 | |
| Throat | 28.9 | 38 | 66.5 | Ser | 34.1 | |
| Throat | 29.1 | 36.8 | 66.5 | Ser | 34.4 | |
| Throat | 29.5 | 36.3 | 66.5 | Ser | 34 | |
| Urethra | - | - | no peaks | | N/A | |
| Urethra | - | - | no peaks | | N/A | |
| Anal | - | - | no peaks | | N/A | |
| Anal | - | - | no peaks | | N/A | |
| Anal | - | - | no peaks | | N/A | |
| Anal | - | - | no peaks | | N/A | |
| Urethra | - | - | no peaks | | N/A | |
| Anal | - | - | no peaks | | N/A | |
| Anal | - | - | no peaks | | N/A | |
| Throat | - | - | no peaks | | N/A | |
| Throat | - | - | no peaks | | N/A | |
| Anal | - | - | no peaks | | N/A | |
| Anal | - | - | no peaks | | N/A | |
| Throat | - | - | no peaks | | N/A | |
| Anal | - | - | no peaks | | N/A | |
| Anal | - | - | no peaks | | N/A | |
